# Immune cell residency in the nasal mucosa and COVID-19 severity across the age range

**DOI:** 10.1101/2021.02.05.21251067

**Authors:** Konner Winkley, Dithi Banerjee, Daniel Louiselle, Rebecca Biswell, Nyshele Posey, Kelly Fatheree, Stephanie McDanel, Todd Bradley, Mary E. Moffatt, Boryana Koseva, Warren A Cheung, Jeffrey J Johnston, Rangaraj Selvarangan, Tomi Pastinen, Elin Grundberg

**Affiliations:** Genomic Medicine Center, Children’s Mercy Research Institute, Children’s Mercy Kansas City, Kansas City, MO, 64108; Department of Pathology and Laboratory Medicine, Children’s Mercy Kansas City, Kansas City, MO, 64108; Division of Emergency Medicine, Children’s Mercy Kansas City, Kansas City, MO, 64108

## Abstract

Severe coronavirus disease of 2019 (COVID-19) positively correlates with age (Centers for Disease Control), develops after progression of infection from the upper airway to the lower respiratory tract (LRT), and can worsen into acute respiratory distress syndrome (ARDS) (Shi et al., 2020). Why children seem to be less likely to develop severe disease remains unclear. As the nasal mucosa (NM) is the first site of contact and defense for respiratory pathogens such as SARS-CoV-2 before dissemination to the LRT (Casadei and Salinas, 2019), we hypothesized that differences in this tissue across the age range may help explain the disparity in COVID-19 severity. To this end, we profiled NM samples across the lifespan in health and disease. We find that global transcriptomic changes including the expression of SARS-CoV-2 and coronavirus-associated receptors and factors are not correlated with age or the novel virus type, since pediatric NM cells mount similar antiviral response to both SARS-CoV-2 or Influenza B. Rather, we find immune cell residency in NM decreases dramatically with age especially cells of the innate immune system. This includes a resident-memory-like T cell subset with antiviral properties. These observations give plausible biological explanation to the observed clinical differences in disease spectrum and provide a foundation for future experimental studies.

## Results and Discussion

We characterized the composition of the NM across the lifespan, by taking advantage of readily available clinical samples of NM-derived cells from COVID-19-negative pediatric patients and healthcare workers. We developed a protocol to collect viable cells from salvaged respiratory specimens collected by mid-turbinate swabs shortly after completion of clinical testing. We used a specimen pooling approach, where isolated cells from four to nine deidentified individuals of similar age groups were combined into a common. This increased coverage of cell types in the age-specific pools, and also averaged out the effects of individual heterogeneity, resulting in more accurate population level sampling. Applying single-cell RNA sequencing (scRNAseq), our approach resulted in the transcriptomic profiling of ∼30,000 cells across six pools with average ages of 1, 7, 13, 17, 33, and 50 years which effectively covers the vast majority of the lifespan.

We detected the expected epithelial and immune cell types in our data (Fig 1a). The epithelial cell types included multiciliated cells (Pardo-Saganta et al., 2015), deuterosomal precursor cells (Revinski et al., 2018; Ruiz García et al., 2019), secretory cells (Mori et al., 2015; Pardo-Saganta et al., 2015), basal cells and suprabasal cells (BOERS et al., 1998; Mori et al., 2015). We also classified six distinct immune cell types which were: monocytes, conventional dendritic cells (cDCs), plasmacystoid dendritic cells (pDCs), B cells, Mast cells, and T cells. Additionally, we were able to identify *CFTR* expressing ionocytes (Plasschaert et al., 2018). We confirmed our cell type assignments by comparing marker genes for each cell type to previously published cell type markers of nasal epithelial cells (Chua et al., 2020; Deprez et al., 2020; Ruiz García et al., 2019) (Extended Data Table 1).

**Figure 1:**
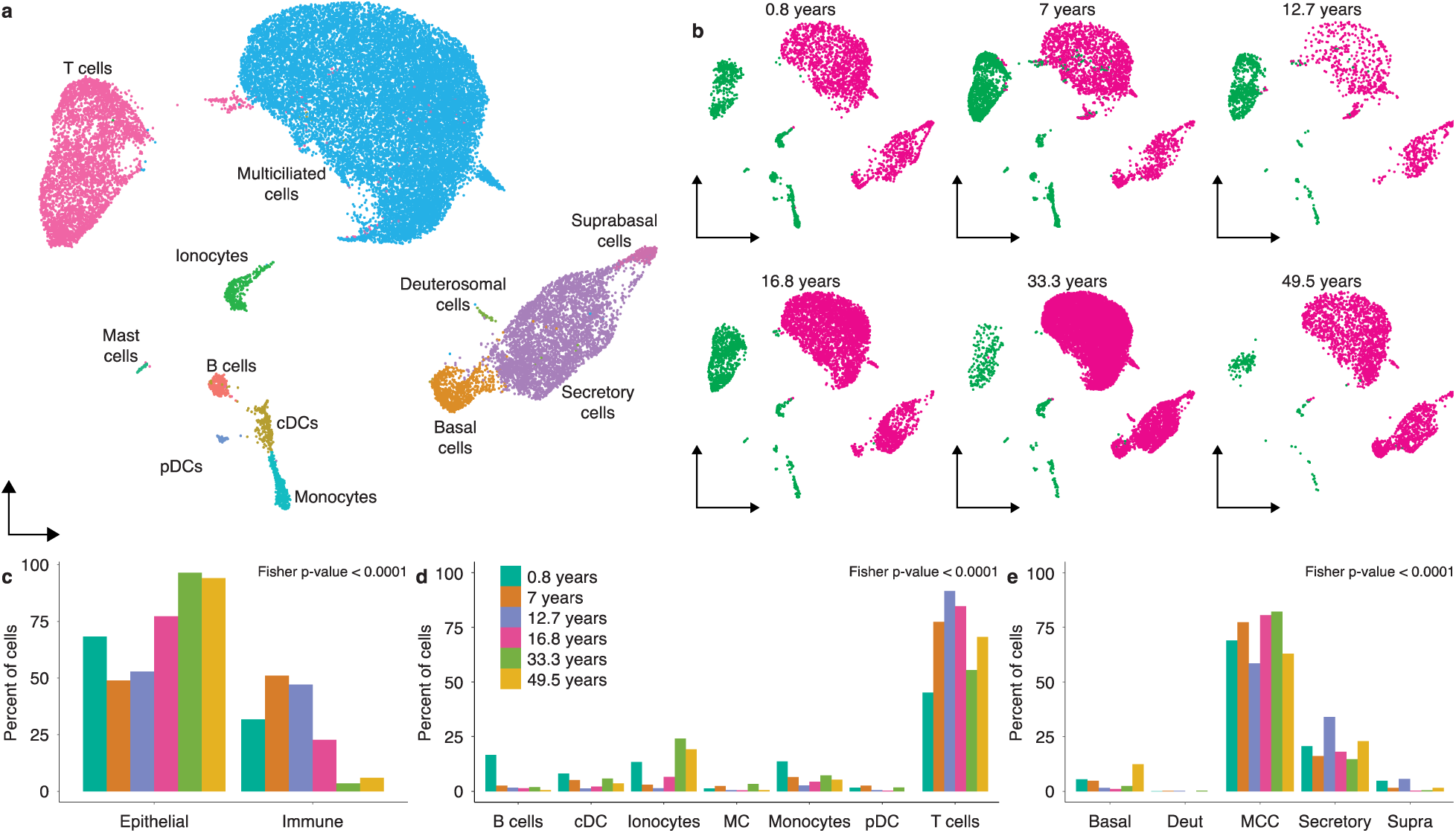
Immune cell residency decreases with age in the nasal mucosa. A) UMAP projection of cells from nasopharyngeal swabs collected across the lifespan pools. Colors correspond to different cell types. B) Same UMAP projection from A split by the 6 pools that make up the dataset. Cells are colored by a coarse cell type assignment where magenta cells are epithelial, and green cells are immune. C) Bar plot quantification of B. D-E) Percent of cells in the immune (D) and epithelial (E) cell subsets that fall into each cell type. Colors in D and E are consistent with C. p-values in C-E are for a Fisher’s exact test of association between age and cell type composition.

We next compared the proportions of cell types across the lifespan and found a striking negative relationship (Fisher test for interaction between age and coarse cell type proportion p-value < 0.0001) between age and immune cell residence in the NM samples which did not seem to be driven by gain or loss of one, or a few, specific cell type (Fig 1b-d). We found that the predominant immune cell type in these NM samples across the age range is a population of T cells (Fig 1d). Many studies investigating the healthy NM with single-cell transcriptomics have only focused on adult samples (Ballestar et al., 2020; Chua et al., 2020; Deprez et al., 2020). Because adults have a relatively small percentage of immune cells comprising their NM, this T cell population has typically lacked the coverage and resolution required for proper characterization. However, because of the increased immune cell residency in adolescents, we were able to capture and identify over 4,500 T cells in our dataset. With this increased resolution, we found that these NM T cells were primarily CD8 positive and expressed genes encoding CCL5 and interferon gamma, TNF, PRF, granzymes, and killer-like receptors that are important for T cell effector function and antiviral activity. Additionally, these cells are strongly enriched for expression of tissue residency markers such as *ITGAE* (CD103), *ITGA1* (CD49a) and *CD69*, and are void of expression of *CCR7*, which is necessary for lymphocyte homing (Fig 2). This transcriptional profile is very similar to that of resident-memory T lymphocytes (T_rm_) (Schenkel and Masopust, 2014). T_rm_ cells are known to occupy other tissues including the lungs in humans; they are important for local immune response there and can serve as a functional link between the innate and adaptative immune system (Bedford et al., 2020; Kurd et al., 2020; Steinbach et al., 2018; Sun et al., 2019). While T_rm_ occupancy in the NM has yet to be described in humans, T_rm_ are known to occupy the NM in mice and help to prevent pulmonary dissemination of influenza virus (Pizzolla et al., 2017) This characterization of NM T cells demonstrates the potential insights gleaned by profiling NM samples across the lifespan.

**Figure 2:**
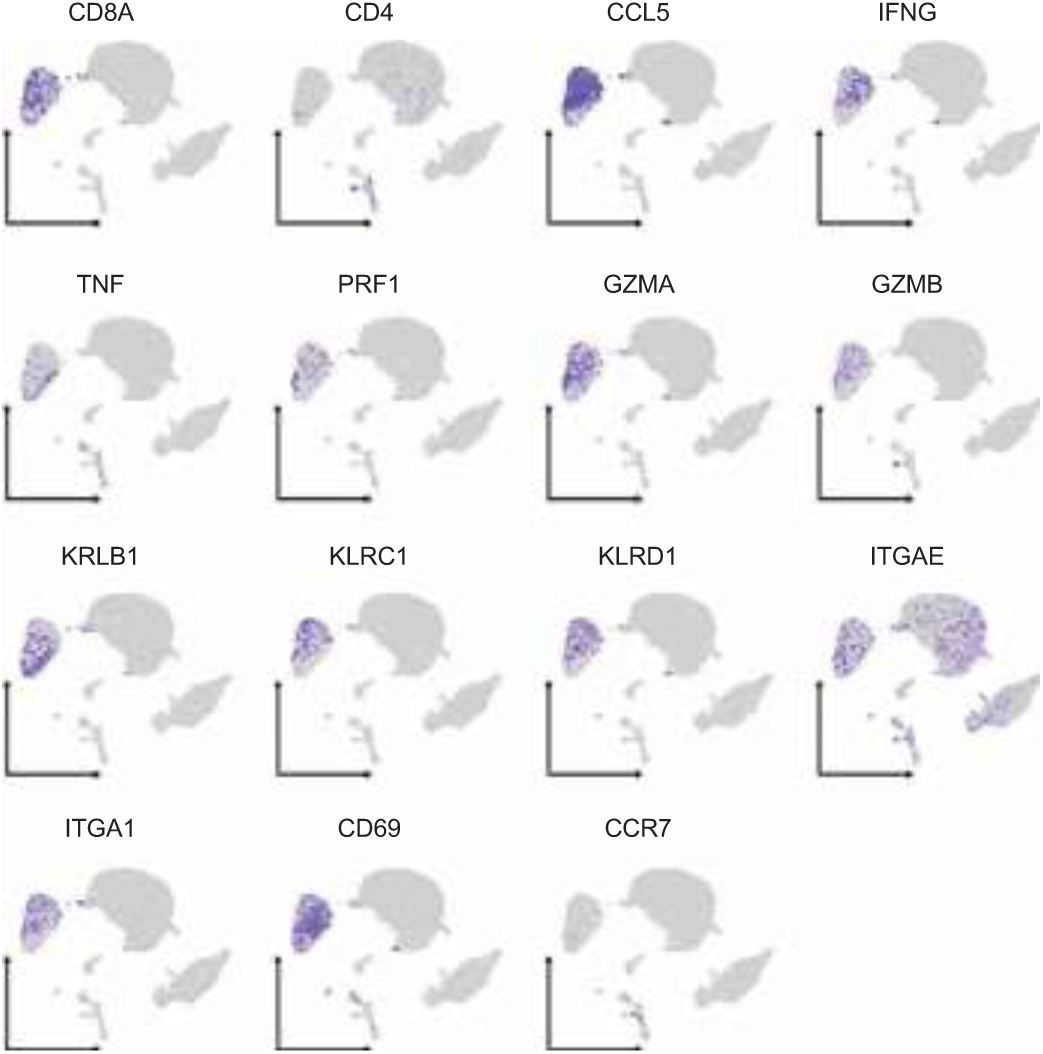
The primary immune cell population in the NM has an anti-viral resident-memory-like T cell transcriptional profile. UMAP projection is identical to Figure 1A. Depth of color for each dot represents level of expression for each specific gene in the given cell.

To validate the observed age-association of immune cell residency we first compared the cellular composition in our 33-year-old pool with a published dataset of healthy individuals whose average age was 36 years (Chua et al., 2020). We found high concordance in the proportions of cell types in this age-matched comparison (Extended Data Fig 1). Next, we performed orthogonal validation by genome-wide epigenome profiling of independent NM samples across the age range (0-45 years). We found that regulatory regions of the genome associated with immune cell activity (Meuleman et al., 2020) were significantly less methylated (ie. hypomethylated) in samples from younger patients (Extended Data Fig 1) suggesting higher transcriptional activity of immune cells in these samples. While the methylation levels were significantly (p < 0.0001) lower across the age range in both myeloid and lymphoid lineages, the changes were more pronounced in the myeloid lineage (Extended Data Fig 2). This further indicates the link between the innate immune system and the observed age-association of NM cell composition.

The observed negative association of immune-cell residency with age likely has an impact on the progression of COVID-19. Another plausible variable affecting disease progression is the expression level of SARS-CoV-2 related genes (SCARFs) (Singh et al., 2020a). However, we found that none of the SCARFs demonstrated a relationship between expression level and age (Fig 3). This included the primary SARS-CoV-2 entry factors, *ACE2* and *TMPRSS2* (Extended Data Fig 3a) (Sungnak et al., 2020; Xu et al., 2020b). These two genes displayed a cell-type specific expression pattern consistent with earlier reports and did not change across the age range (Extended Data Fig 3b-c) (Chua et al., 2020; Deprez et al., 2020; Singh et al., 2020b; Ziegler et al., 2020). Additionally, we explored genome wide expression differences in the NM with respect to age. We found little evidence of fundamental expression changes independent of cell type across the age range that might contribute to the muted severity of COVID-19 in younger individuals (Extended Data Fig 4a-b). From this, we conclude that variation in observed disease severity across the age range are unlikely to be explained by transcriptomic differences, but rather differences in immune cell residency.

**Figure 3:**
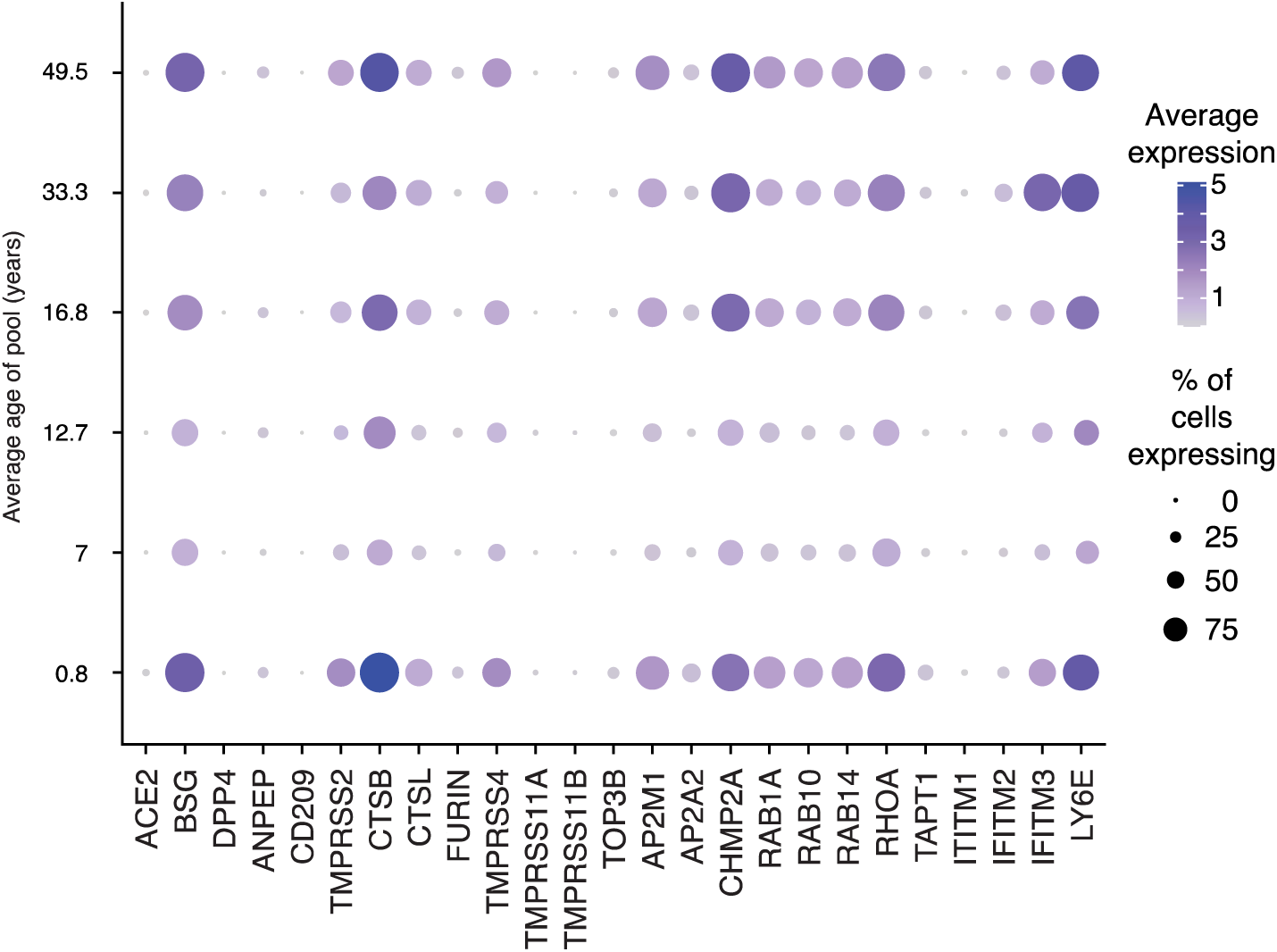
Expression level of SCARFs is not correlated with age. Each row represents the entire population of epithelial cells in Figure 1B. Dot size scales with the percent of cells in the population where the given gene is detected. Dot color shows the average scaled and normalized expression level in the cells where the specific gene is detected. Darker colors are higher in relative expression in the expressing cells.

Next, we hypothesized that the pediatric response to SARS-CoV-2 may be altered in some way that may contribute to mild disease, such as an upregulation of anti-inflammatory molecules, or activation of an immunosuppressive cell type. To test this, we performed scRNAseq on NM samples of three COVID-19 positive pediatric patients (5-11 years). We found that pediatric NM cells were not unexpectedly refractory to SARS-CoV-2 infection (Extended Data Fig 5), and that pediatric COVID-19 positive patients did not display increased immune cell infiltration of the NM (Fig 4a-b). We tested for activity of immune cells by quantifying cell-cell interactions via expression patterns of ligand-receptor pairs (Efremova et al., 2020). We observed a slight increase in epithelial-to-immune interactions in COVID-19 positive children compared to COVID-19 negative children especially in T cells and monocytes (Extended Data Fig 6a) further highlighting the role of the innate immune system. However, these interactions did not result in marked increase in cell death of epithelial cells (Extended Data Fig 6b). This is in stark contrast to the NM immune response in adults with moderate and severe COVID-19 where dramatic infiltration of immune cells and resulting epithelial cell death are observed. However, in line with other reports, we did note a strong upregulation of interferon signaling in the NM of COVID-19 positive pediatric patients (Fig 4c).

**Figure 4:**
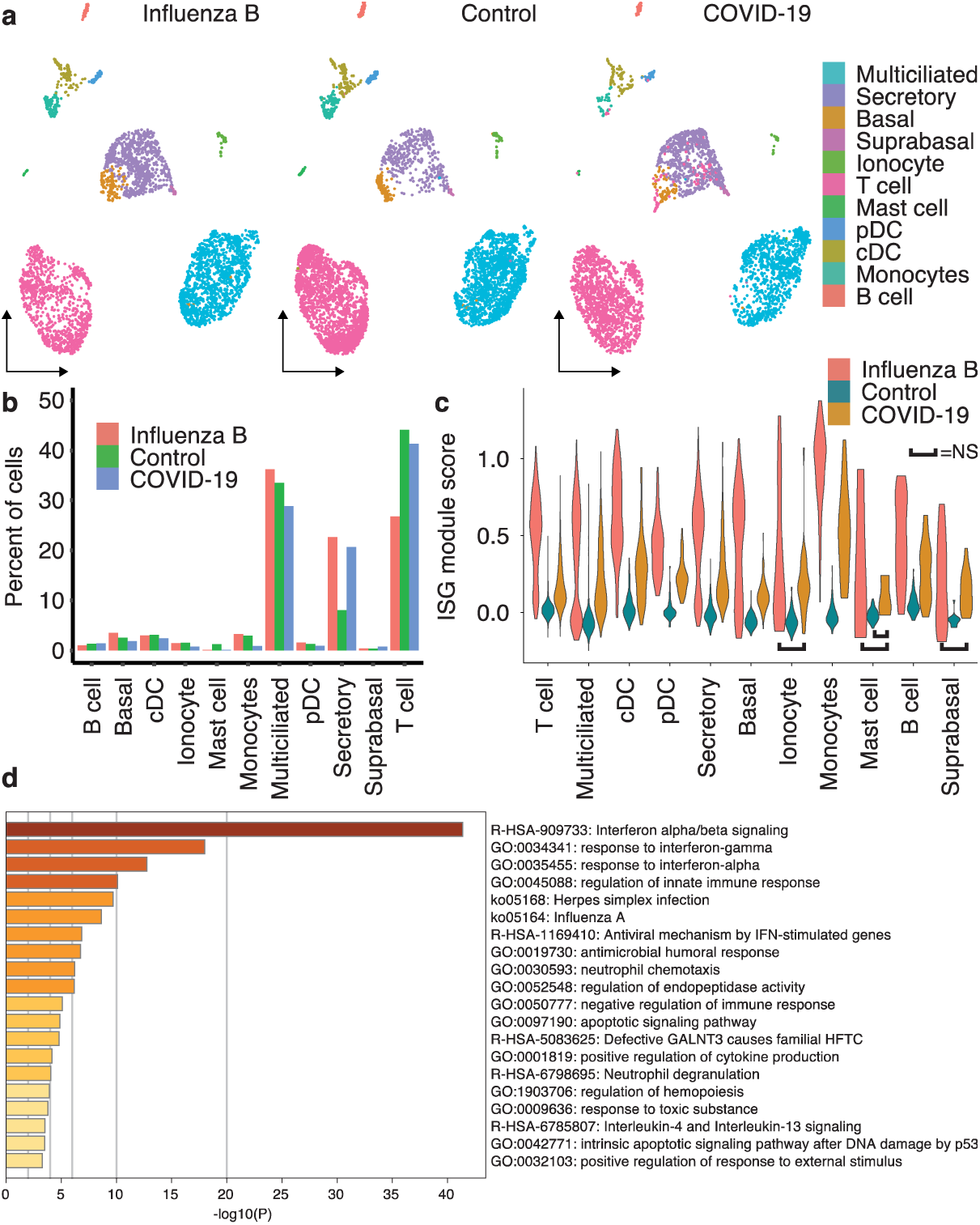
Cellular response to mild cases of COVID-19 in pediatric patients is similar to influenza B and is characterized by increased interferon signaling without increased immune cell infiltration. A) UMAP projections of cells from nasal and nasopharyngeal swabs of pediatric patients. Control group is the same as the 7-year-old pool in Figure 1B. UMAP space across all three groups is identical. B) Bar plot quantification of the cell types present in A. C) Violin plots representing the interferon stimulated gene module score of each cell in each cell type separated by infection status. All pairwise comparisons of viral state within a cell type are significant with a p-value of <0.0001 unless marked which indicates a p-value > 0.05. D) Metascape analysis of most highly enriched terms for the set of genes that is at least Log_2_ fold upregulated in both Influenza B positive samples and in COVID-19 positive samples compared to control samples.

To further characterize the immune response to SARS-CoV-2 in children versus adults, we expanded our COVID-19 pediatric cell atlas by including peripheral blood mononuclear cells of these same COVID-19 positive children. We found that the peripheral immune response to COVID-19 infection is quite similar to local NM immune response. There are no significant expansions of specific immune cell types in the periphery (Extended Data Fig 7a-b), in contrast to what has been observed in the circulating leucocytes of infected adults (Chua et al., 2020; Tang et al., 2020; Wilk et al., 2020; Xu et al., 2020a). Additionally, the transcriptional response in the periphery is primarily marked by increased interferon signaling (Extended Data Fig 7c). This leads us to conclude that the pediatric immune response to COVID-19 is not altered in an unexpected and immunosuppressive way that results in more mild disease severity. Instead, both the local and peripheral immune systems display the hallmarks of antiviral and interferon signaling in response to viral infection.

Finally, we asked if this anti-viral response in the NM to COVID-19 infection in pediatric patients is unique by nature of the novel Coronavirus. To test this, we contrasted our pediatric COVID-19 NM profiles with age- and tissue-matched scRNAseq profiles derived from Influenza B infected children. We found that the NM immune response to COVID-19 is strikingly similar to the response to influenza B infection in pediatric patients (Fig 4a-c & Extended Data Fig 6) and is primarily characterized by robust interferon signaling. (Fig 4d).

In conclusion, we present the first-of-its kind single-cell atlas that spans the dimensions of age, tissue, and virus types with important and novel coverage during childhood. Using this resource, we provide important insight into underlying factors for the observed age-association of COVID-19 disease severity during the current pandemic. We demonstrate how this resource can be used to drive future experimental studies. For example, we identify the T cell population in the NM as having a T_rm_-like transcriptional profile. While the implications of this population remain to be fully explained, there is growing evidence that cross-reactive T cells to endemic human coronavirus (CoVs) exist (Braun et al., 2020; Hu et al., 2020; Lee et al., 2020; Stervbo et al., 2020), and it is established that children tend to have higher burden of these endemic CoVs infections compared to adults (Monto et al., 2020). Therefore, one could hypothesize that NM T_rm_ may represent a reservoir of potential cross-reactive T-cells that offer protection from SARS-CoV-2 infection. Other studies will be needed to fully test this hypothesis, and our atlas provides a foundation for them.

While our pooling strategy allowed us to obtain sufficient number of high-quality cells for cataloging all major cell populations it limited our ability for high-powered interindividual analysis of minor differences. Additionally, possible confounding factors including racial differences among our study populations, and the effect of non-SARS-CoV-2 respiratory viruses could not be fully explored. However, we performed orthogonal and multi-dataset external validation whenever possible to try and identify if these factors created a major confounding effect. In closing, our initial analyses of this resource raise the possibility that pediatric COVID-19 severity may be less because child and adolescent NM may be inherently primed for immune response to SARS-CoV-2 and not because of unexpected transcriptional or immune response differences across the age range. The increased presence and activity of resident immune cells in the NM of children and adolescents may be capable of clearing SARS-CoV-2 infection without dissemination to the LRT, and subsequent development of ARDS.

## STAR Methods

### Patient Recruitment

All study subjects were enrolled at Children’s Mercy either using salvage sample collection protocol or using prospective cohort study protocols. Specifically, the NM cohort of COVID-19 negative individuals included patients aged 0 to 18 years tested for COVID-19 as a part of their standard of care procedure, and healthcare workers screened for COVID-19, these samples were collected using a salvage sample protocol (IRB # STUDY00001258). Similarly, patients undergoing multiplex testing for respiratory viruses were regularly screened and all children positive for Influenza B were selected for the NM cohort of Influenza B positive children these samples were collected using salvage sample protocol (IRB # STUDY00001193). For both groups, subjects were deidentified and samples stored at 4°C were collected within 12 hours of test results.

For COVID-19 positive children, families were enrolled in the CODIEFY study approved by the Institute Review Board (IRB) at Children’s Mercy (IRB # STUDY00001317). Parents or legally appointed representatives of COVID-19 positive children were approached for enrollment and verbal consent within 24-48 hours of their test results, and children aged 7 years and above have given verbal informed assent. Respiratory and blood specimens were collected and transported by a home-health care nurse following standard precautions within the next 24-48 hours. Samples were processed for nasal cell isolation and PBMC isolation within 2 - 4 hours of collection.

Finally, the PBMC cohort of COVID-19 negative children were enrolled in the Genomics Answers for Kids Program approved by the Institute Review Board (IRB) at Children’s Mercy (IRB # 11120514). Participants have given written informed consent to collect pertinent medical information and family history from their medical record and give a blood sample. Blood draws for the study were paired with clinical draws when possible.

### Mid-turbinate swab sample collection

Selected patients were screened for either COVID-19 or Influenza by mid-turbinate swab at Children’s Mercy Kansas City. Respiratory samples were collected using Copan FLOQSwabs (Copan Cat No. 56780CS01 for patients less than 2 years old or Copan Cat No. 56380CS01 for patients greater than 2 years old). Samples were stored at 4°C in BD Universal Transport Media (BD Cat No. 220220) until transported to the laboratory. Experimental sample processing began within 12 hours of sample collection after confirmation of positive or negative test for COVID-19 or Influenza B via RT-PCR.

### Cell collection from mid-turbinate swabs and cryopreservation

Each sample was diluted with cold PBS (Thermo Fisher Cat No. 14190144) + 2% FBS (GE Healthcare Cat No. SH30088.03HI) up to a total volume of 5 mL and passed through a 40-µm nylon mesh cell strainer that had been prewetted with 2 mL of PBS + 2% FBS. The strainer was then rinsed with 7 mL of cold PBS + 2% FBS. The sample was transferred to a 15-mL conical tube and centrifuged at 300 x *g* at 4°C for 8 minutes. The supernatant was carefully removed without disturbing the cell pellet. The cell pellet was resuspended in 200 µL of cold PBS + 2% FBS, and the cell count and viability were assessed using 0.4% Trypan Blue and a Countess II automated cell counter. The cell suspension was transferred to a 1.5-mL tube and centrifuged at 300 x *g* at 4°C for 8 minutes, and the supernatant was carefully removed without disturbing the cell pellet. The cell pellet was resuspended in 1 mL of cold Recovery Cell Culture Freezing Medium (Thermo Fisher Cat No. 12648010), and the cell suspension was transferred to a cryogenic storage vial. The cryogenic storage vial was placed in a Corning CoolCell FTS30, which was then placed in a −80°C freezer overnight. Samples were stored at −80°C for no longer than one week before being thawed and processed for scRNAseq.

### Cell pooling and single-cell RNA-sequencing of NM

Samples with less than 30% viability were excluded from analysis and cells were used in pools or individually. For the lifespan atlas, six groups were formed based on the age of the patients: Control Group 1) 4 months – 18 months (n=7); Control Group 2) 5 years – 9 years (n=5); Control Group 3) 11 years – 15 years (n=6); Control Group 4) 16 years – 19 years (n=9); Control Group 5) 30 years – 35 years (n=4); and Control Group 6) 36+ years (n=4). For the Influenza B positive samples, a single pool of four samples was created (4-11 years, n=4). COVID-19 positive samples (5-11 years, n=3) were processed individually. To reduce cell stress caused by delays due to processing multiple samples in parallel, the 6 lifespan pools were processed in 2 batches of 3 groups each. Samples within the first batch were thawed and pooled as described below and carried through to the first stable pause point in the scRNAseq protocol (GEM-RT Incubation); the process was then repeated for the second batch of samples. Once both batches of samples had reached the first pause point, all samples were processed in parallel to completion. Influenza B positive samples were processed independently.

For each sample to be thawed, 10 mL of Thawing Medium consisting of DMEM/F-12 (Thermo Fisher Cat No. 11320033) supplemented with 10% FBS and 100 units/mL of penicillin and 100 µg/mL of streptomycin (Thermo Fisher Cat No. 15140122) was prewarmed in a 37°C bead bath. Each cryogenic storage vial containing a sample to be thawed was placed in the 37°C bead bath. No more than 5 samples were thawed at a time. When only a small ice crystal remained in the sample, both the cryogenic storage vial and the 15-mL conical tube containing the Thawing Medium were aseptically transferred to the biosafety cabinet. 1 mL of Thawing Medium was slowly added, dropwise, to the sample. The diluted sample was then mixed gently by pipetting and further diluted in the remaining 9 mL of Thawing Medium. The thawed and diluted cells were left at room temperature while the remaining samples were similarly thawed. When all samples in the batch were thawed, the samples were centrifuged at 300 x *g* for 8 min. The supernatant was carefully removed without disturbing the cell pellets. The cell pellets were each resuspended in 0.5 mL of Thawing Medium, and the cell suspensions were placed on ice. The cell suspensions were then combined and pooled together in age-defined groups. Each resulting pool was passed through a prewetted 40-µm nylon mesh cell strainer, and the cell strainers were rinsed with 5 mL of cold Thawing Medium. The pooled cell suspensions were centrifuged at 300 x *g* for 8 min at 4°C, and the supernatant was carefully aspirated without disturbing the cell pellets. The cell pellets were resuspended in 100 µL of cold Thawing Medium, and cell count and viability were assessed using 0.4% Trypan Blue and a Countess II automated cell counter. For each control group, 4 wells of a Chromium Chip B (10x Genomics Cat No. 1000153) were loaded with 20,000 cells each; for each Influenza B group, 2 wells of a Chromium Chip B (10x Genomics Cat No. 1000153) were loaded with 32,000 cells each; for each SARS-CoV-2 positive sample, 2 wells of a Chromium Chip B (10x Genomics Cat No. 1000153) were loaded with 16,000 cells each. Following cell loading, scRNAseq was performed identically for all samples using the Chromium Single Cell 3’ Library & Gel Bead Kit v3 (10x Genomics Cat No. 1000075) according to the manufacturer’s protocol.

### PBMC collection and single-cell RNA sequencing of PBMC

Blood was collected in sodium heparin tubes and stored at 4°C until they were de-identified and processed. PBMCs were isolated using the EasySep Direct Human PBMC Isolation Kit (STEMCELL Technologies Cat No. 19654RF) on a STEMCELL Technologies RoboSep-S automated platform according to the manufacturer’s protocol. Following isolation, the cell suspension was centrifuged at 300 x g for 8 minutes, and the supernatant was carefully aspirated. Residual RBCs were lysed by resuspending the cell pellet in 1 mL of ACK Lysing Buffer (Thermo Fisher Cat No. A1049201) and incubating at room temperature for 5 minutes. The cell suspension was centrifuged at 300 x g for 8 minutes, and the cell pellet was washed twice with PBS (Thermo Fisher Cat No. 14190144) supplemented with 2% heat-inactivated FBS (GE Healthcare Cat No. SH30088.03HI). Cell count and viability were assessed using a Countess II automated cell counter. Cells were cryopreserved in aliquots of at least one 1 × 10^6^ cells by centrifuging the cells at 300 x g for 8 minutes, aspirating the supernatant, and resuspending the cell pellets in Recovery Cell Culture Freezing Medium (Thermo Fisher Cat No. 12648010). Cells were frozen at a rate of −1°C/minute in a Corning CoolCell FTS30 and stored at −80°C for up to several weeks before they were thawed.

For each sample to be thawed, 10 mL of Thawing Medium consisting of IMDM (ATCC Cat No. 30-2005) supplemented with 10% heat-inactivated FBS (GE Healthcare Cat No. SH30088.03HI), 100 units/mL of penicillin, and 100 µg/mL of streptomycin (Thermo Fisher Cat No. 15140122) was prewarmed in a 37°C bead bath. When each cryopreserved sample was thawed and diluted, as described above for the nasal samples, they were centrifuged at 300 x *g* for 8 min, the supernatant was carefully aspirated, and the cell pellets were resuspended in 0.5 mL of room-temperature Thawing Medium. COVID-19 positive samples were processed individually, and age-matched controls were processed in pools of 32 samples/pool. For the pools, up to 1 million cells from each sample were pooled together on ice, and the resulting pool was passed through a 40-µm nylon mesh cell strainer to remove cell aggregates. Cell count and viability of all samples were assessed using a Countess II automated cell counter. For each control pool, 4 wells of a Chromium Chip B (10x Genomics Cat No. 1000153) were loaded with 25,000 cells each, and for each SARS-CoV-2 positive sample, 2 wells of a Chromium Chip B (10x Genomics Cat No. 1000153) were loaded with 16,000 cells each. Following cell loading, scRNAseq was performed identically for all samples using the Chromium Single Cell 3’ Library & Gel Bead Kit v3 (10x Genomics Cat No. 1000075) according to the manufacturer’s protocol.

### Specimen Pooling and DNA Isolation for methylation samples

Nasal specimens were stored at −80°C and were brought to room temperature before pooling by age. Before pooling, the specimens were mixed well with gentle pipetting. 100µL from each specimen was removed and pooled together in a 1.5mL tube. Once all specimen aliquots were added to the pool, the pool was mixed by pipetting and 200μl was taken from each pool into a new 1.5mL tube for DNA isolation. DNA was isolated with a DNeasy Blood and Tissue Kit (Qiagen, Cat No. 69504) with the following modifications to kit protocol: 8uL of RNase A was used instead of 4ul during the optional RNase A step and the lysis incubation time at 56°C was increased to at least 3 hours to ensure complete lysis of the specimens. After isolation, the DNA concentration of each sample was determined using a Qubit dsDNA HS Assay Kit (Fisher, Cat No. Q32851).

### WGBS Library Preparation

100ng of DNA was aliquoted from each sample. Unmethylated λDNA was added to each sample at 0.5%w/v and the samples were sheared mechanically using a Covaris LE220-plus system to a length of 350 bp, using the settings recommended by the manufacturer. The sizing was determined by a High Sensitivity D1000 ScreenTape and Reagents (Agilent, Cat. No. 5067-5584 and 5067-5585) on the TapeStation platform. Once the input DNA was at the proper fragment size, the samples were concentrated with a SpeedVac to a volume of 20µL. The samples then underwent bisulfite conversion with an EZ DNA Methylation-Gold kit (Zymo, Cat. No. D5006). The samples were eluted off the spin columns with 15μl of low EDTA TE buffer (Swift, Cat. No. 30024) before library preparation.

The low-input libraries were prepared using an ACCEL-NGS Methyl-Seq Library kit (Swift, Cat. No. 30024) with a Methl-Seq Set A Indexing Kit (Swift, Cat. No. 36024), following the protocol associated with the library kit. During the protocol, bead cleanup steps were performed with SPRIselect beads (Beckman Coulter, Cat. No. B23318). Following the recommendation of the kit, 6 PCR cycles were performed to amplify the samples. The final libraries were quantified with a Qubit dsDNA HS Assay Kit and the size was determined by using a BioAnalyzer High Sensitivity DNA Kit (Agilent, Cat. No. 5067-4626).

### Sequencing

Sequencing was performed using an Illumina NovaSeq 6000. Runs of WGBS were 2×151 cycle paired-end, while runs of scRNAseq were 2×94 cycle paired-end.

### Post-sequencing analysis scRNAseq

Sequenced reads were initially processed by the cellranger pipeline (v3.1.0) which includes fastq creation, read alignment, gene counting, and cell calling. All samples were mapped to the cellranger GRCh38 v1.2.0 genome. For SARS-CoV-2 viral transcript detection, a custom reference was made using the cellranger GRCh38 v1.2.0 genome and the SARS-CoV-2 genome as described previously (Chua et al., 2020). The resulting cell by gene matrix from the cellranger “count” step was then processed using standard workflows in Seurat (Butler et al., 2018; Stuart et al., 2019). In brief, low quality cells were removed by filtering out cells with a unique gene count lower than 750 and more than 50% mitochondrial reads. The gene counts for remaining cells that passed quality control were then normalized using SCTransform (Hafemeister and Satija, 2019) with the replicate captures as a batch variable. For the age-range comparisons, the 6 age pools were integrated using the FindIntegrationAnchors and IntegratedData functions in Seurat with default parameters. A similar approach was used for the COVID-19 positive and Influenza B positive samples, where they were normalized independently, and integrated with the control samples. The integrated data was then used for linear and non-dimensional reduction, nearest neighbor finding, and unsupervised clustering. Cell types were assigned by examining expression of known genes in the unsupervised clusters, as well as examining markers of the clusters identified using the FindAllMarkers function in Seurat with default parameters.

### Post sequencing analysis WGBS

Sequenced reads were initially processed by the DRAGEN (Edico/Illumina) pipeline (v.1.1.5). Following DRAGEN alignment, duplicate reads were marked with Picard tools (2019) MarkDuplicates and subsequently removed. Bismark (Krueger and Andrews, 2011) was used to assign the methylation ratio at each CpG site. CpG sites were filtered to remove sites that had less than 10x coverage, overlapped ENCODE problematic regions, or overlapped sites in dbSNP as previously described (Busche et al., 2015).

### Genome-wide age correlated expression

To identify pathways which were affected by age at a genome wide scale, we used a hurdle model (Zeileis et al., 2008) to predict the effect of age on gene expression independently of cell type. We used cell type as the predictor for the zero model with a binomal distribution, and both age and cell type as the predictors for the counts model with a negative binomial distribution. We identified genes where age had a significant effect (p-value < 0.01) in the counts model and selected the top and bottom 250 genes when sorted by the effect of age in the counts model. These 250 genes were then input to the Metascape program (Zhou et al., 2019) using the default express analysis parameters.

### Methylation profile analysis

The immune cell composition of the NM was inferred from the methylation profile of samples that were independent of the single-cell RNA-sequencing samples. We measured CpG methylation in regions that have been identified to be primarily accessible in lymphoid and myeloid cells through large scale DNase 1 hypersensitive sequencing and analysis (Meuleman et al., 2020). The regions that were identified as invariantly accessible across cell types were used as a control to test for overall methylation differences. We compared average methylation of CpGs in these regions across the age range and tested for overall shifts in the distributions with a Kruskal-Wallis test. In each case, the result was significant and all pairwise comparisons were made using a Mann-Whitney U test.

### Cell-cell interaction quantification

We quantified the interactions between different cell types in the nasal epithelia of pediatric patients using the cellphoneDB program (Efremova et al., 2020). The SCTransform corrected gene expression values for each infection state were independently input into cellphoneDB using the “statistical analysis” pipeline. The resulting cell-type by cell-type matrix of statistically significant interactions for each infection state were then plotted as clustered heatmaps, using a consistent color scale between the different infection states.

### Gene expression module scoring

Gene expression modules for Interferon stimulated genes and cell death markers were scored using the AddModuleScore function in Seurat. The genes used for each of these modules can be found in Extended Data Table 2.

### Cell-type composition comparison of adult samples

The cellular composition of adult NM samples was compared to with that of healthy age-matched controls from independent data (Chua et al., 2020) by filtering cells with identical QC metrics, scaling both datasets independently with SCTransform and integrating the data using standard Seurat methods. The resulting clusters were then assigned a cell type by manual curation of marker genes. Finally, cellular composition of the samples was calculated and compared.

### Statistics

Differences in expression values between groups of cells were determined using the FindMarkers function in Seurat with default parameters. Tests for differences in gene expression module scores were performed as two-way, unpaired two-sample t tests. Cells are defined as “expressing” a given gene if at least one UMI mapping to the gene was detected in that cell.

## Data Availability

Raw and processed data from all pooled samples is available via the Gene Expression Omnibus (accession number: GSE162864). Raw data from individual samples are available upon request through EGA and dbGAP upon approval from the Data Access Committee. Fully processed data are available for exploration through the UCSC cell browser (lifespan-nasal-atlas.cells.ucsc.edu)

## Author Contributions

Conceptualization: TB, MM, RS, TP, EG

Sample collection: KF, SM

Clinical coordination: DB, KF RS

Laboratory analysis: DL, RB, NP

Data Analysis: JJ, BK, WC, KW

Writing – Original Draft: KW

Writing – Review & Editing: All

Funding acquisition: RS, TP, EG

## Acknowledgements

The contents are solely the responsibility of the authors and do not necessarily represent the official views of the NIH. E.G. holds the Roberta D. Harding & William F. Bradley, Jr. Endowed Chair in Genomic Research and T.P. holds the Dee Lyons/Missouri Endowed Chair in Pediatric Genomic Medicine.

---

**Extended Data Figure 1:**
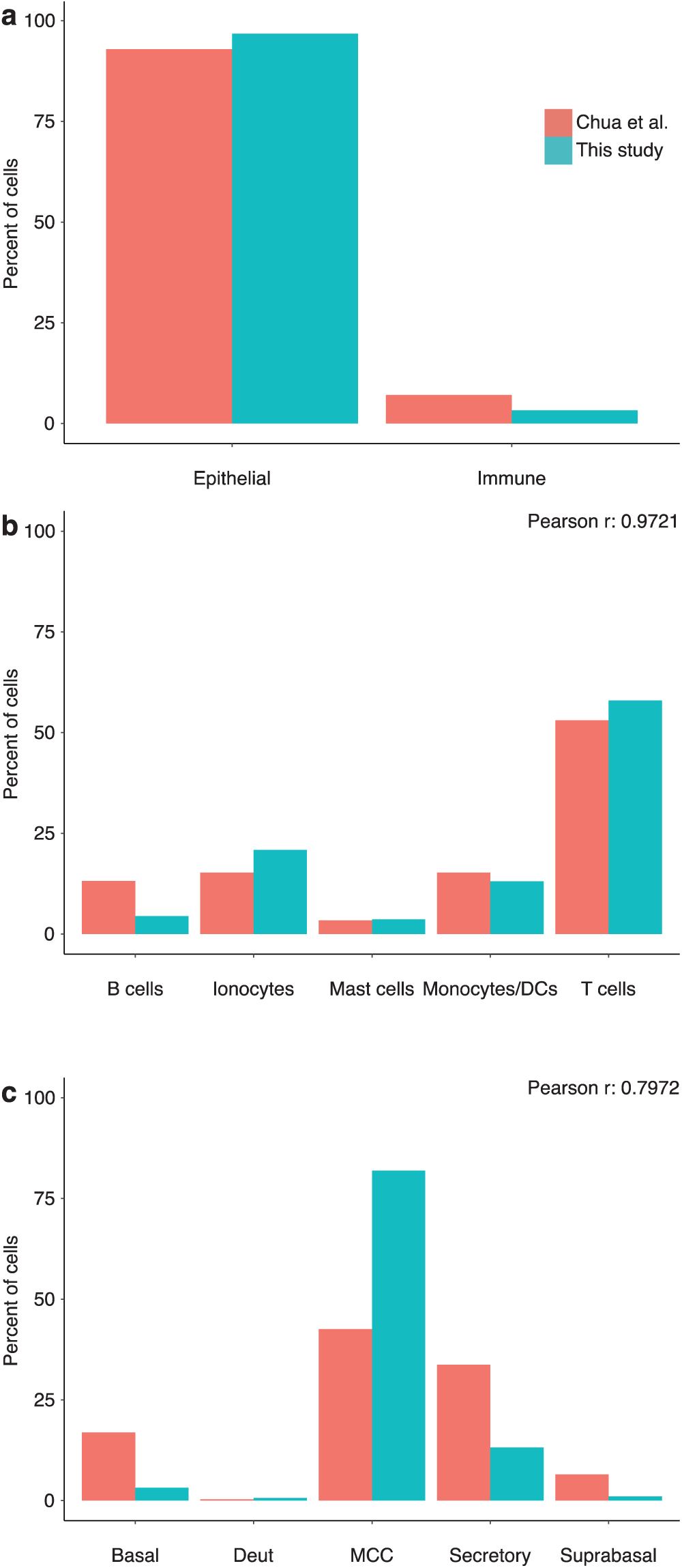
Age-matched cell type proportions in the NM are consistent with other datasets. A) Comparison of epithelial and immune cell proportions in the NM of adults identified by scRNAseq. B-C) Percent of cells in the immune (B) and epithelial (C) cell subsets that fall into each cell type. Average pool age in years: “This study” – 33.3, “Chua et al.” – 35.85.

**Extended Data Figure 2:**
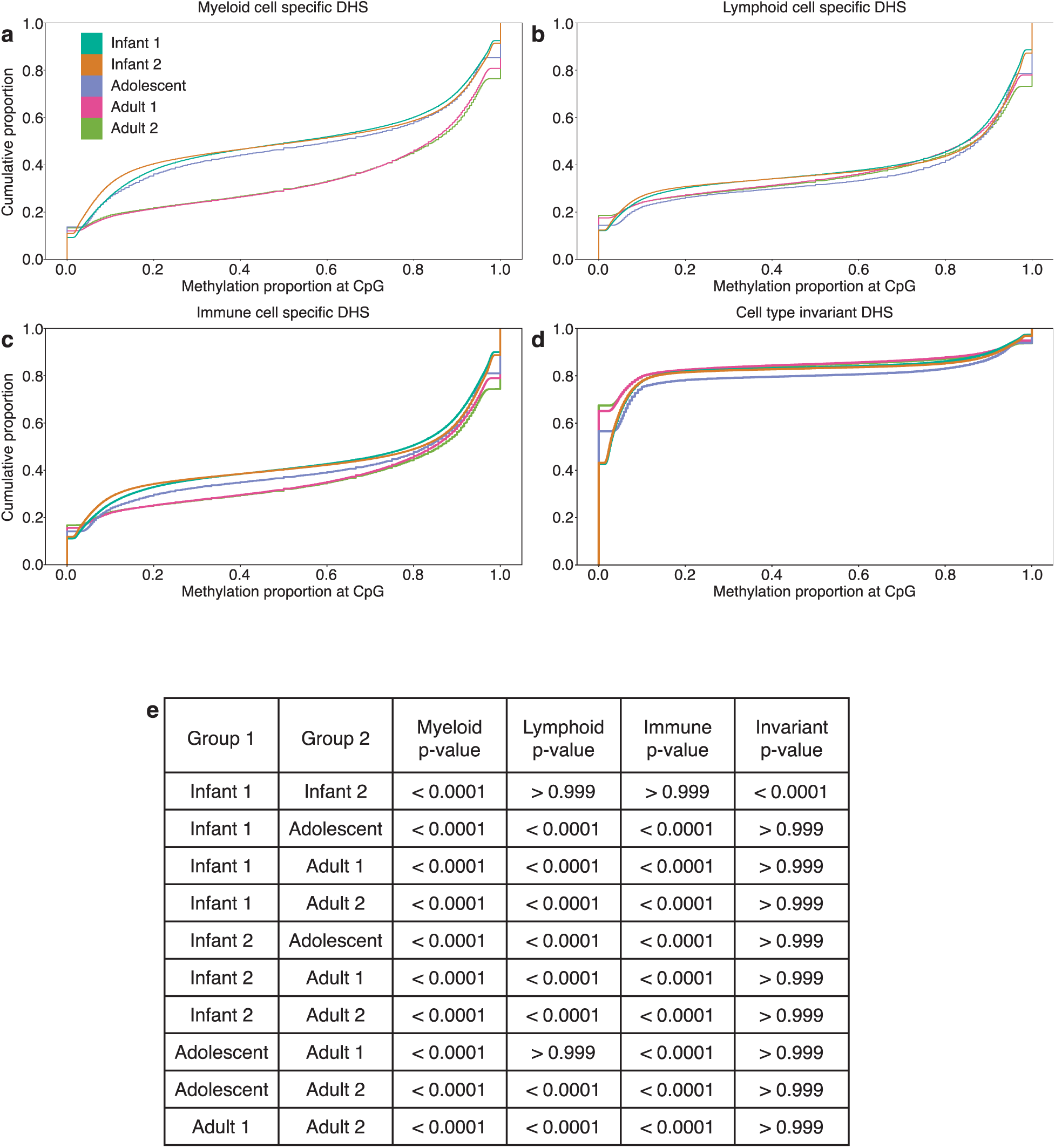
Epigenomic analyses supports the hypothesis that immune residence in the NM decreases with age. A-D) Empirical cumulative distribution function of CpG methylation for all CpGs that overlap DHS sites identified as being enriched in (A) myeloid cells, (B) lymphoid cells, (C) immune (myeloid + lymphoid) cells, and (D) DHS sites that are invariant across cell types. E) Mann-Whitney rank test p-value for the hypothesis that the median methylation ratio of CpG sites overlapping the specified DHS peaks of Group 1 is less than that of Group 2. Average pool age in years: Infant 1 – 0.183, Infant 2 – 0.55, Adolescent – 5.275, Adult 1 – 29.125, Adult 2 – 40.57.

**Extended Data Figure 3:**
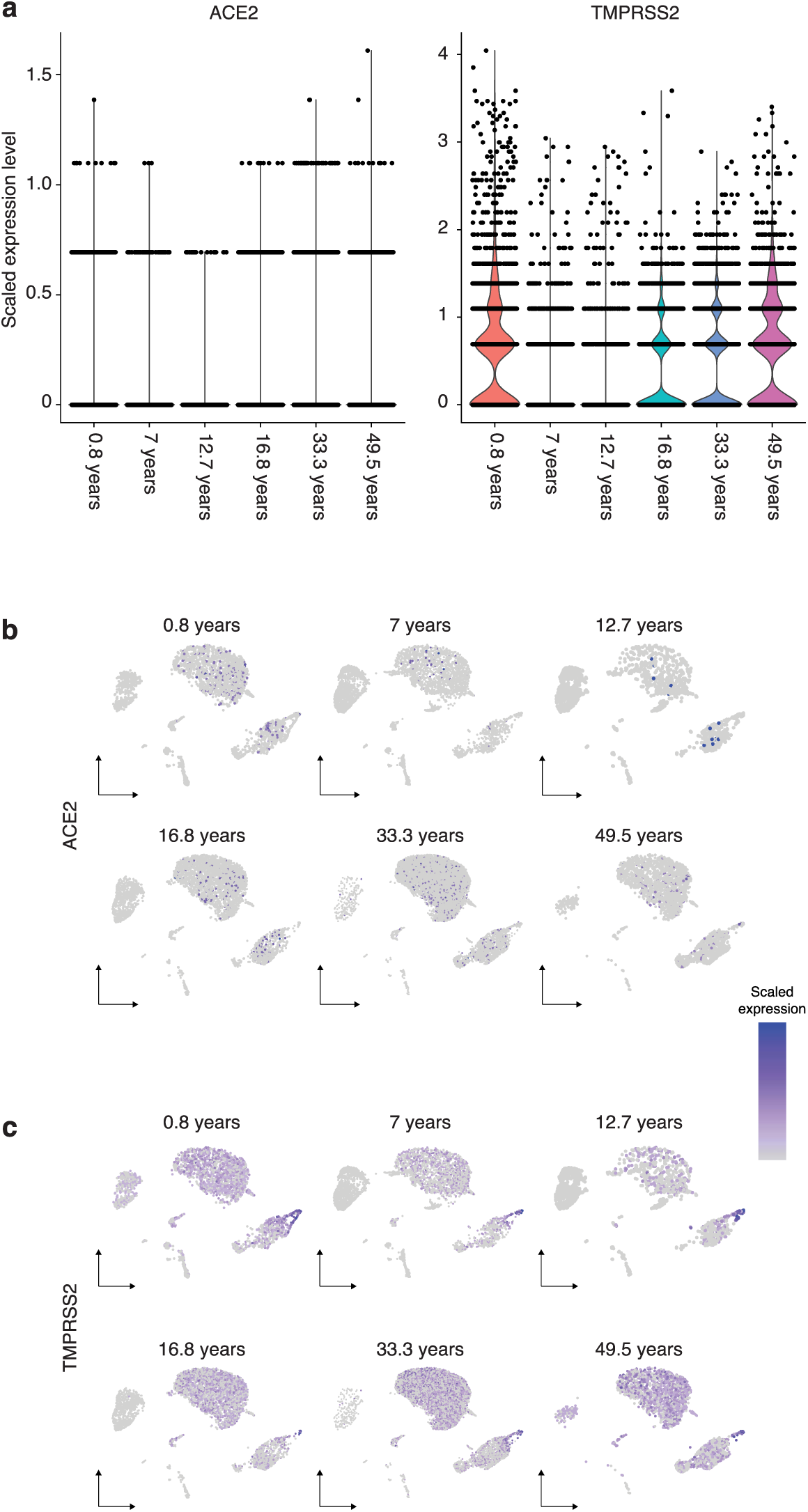
Expression of SARS-CoV-2 entry factors do not change in expression levels, or in their cellular expression pattern across the age range. A) Normalized, scaled gene expression values of ACE2 (left) and TMPRSS2 (right) are similar across the age range. B-C) Cellular expression patterns of ACE2 (B), and TMPRSS2 (C) do not change appreciably across the age range and are consistent with patterns described in previously published reports.

**Extended Data Figure 4:**
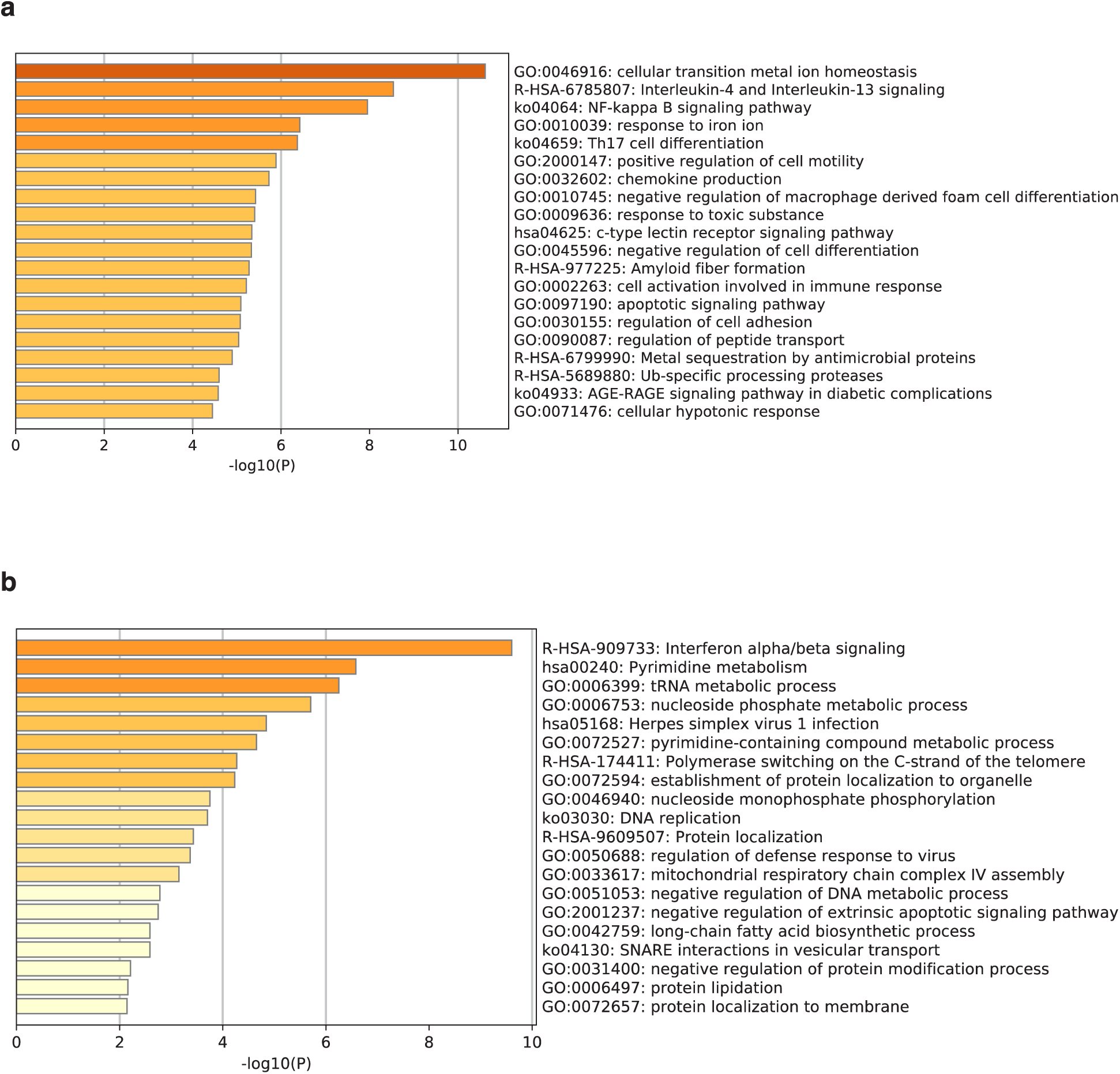
Global transcriptome analysis across the age range does not reveal patterns that explain COVID-19 severity. A-B) Metascape enriched terms for the top 250 statistically significant genes in a hurdle model of which age had the largest positive negative effect on expression (A), or the largest positive effect on expression (B).

**Extended Data Figure 5:**
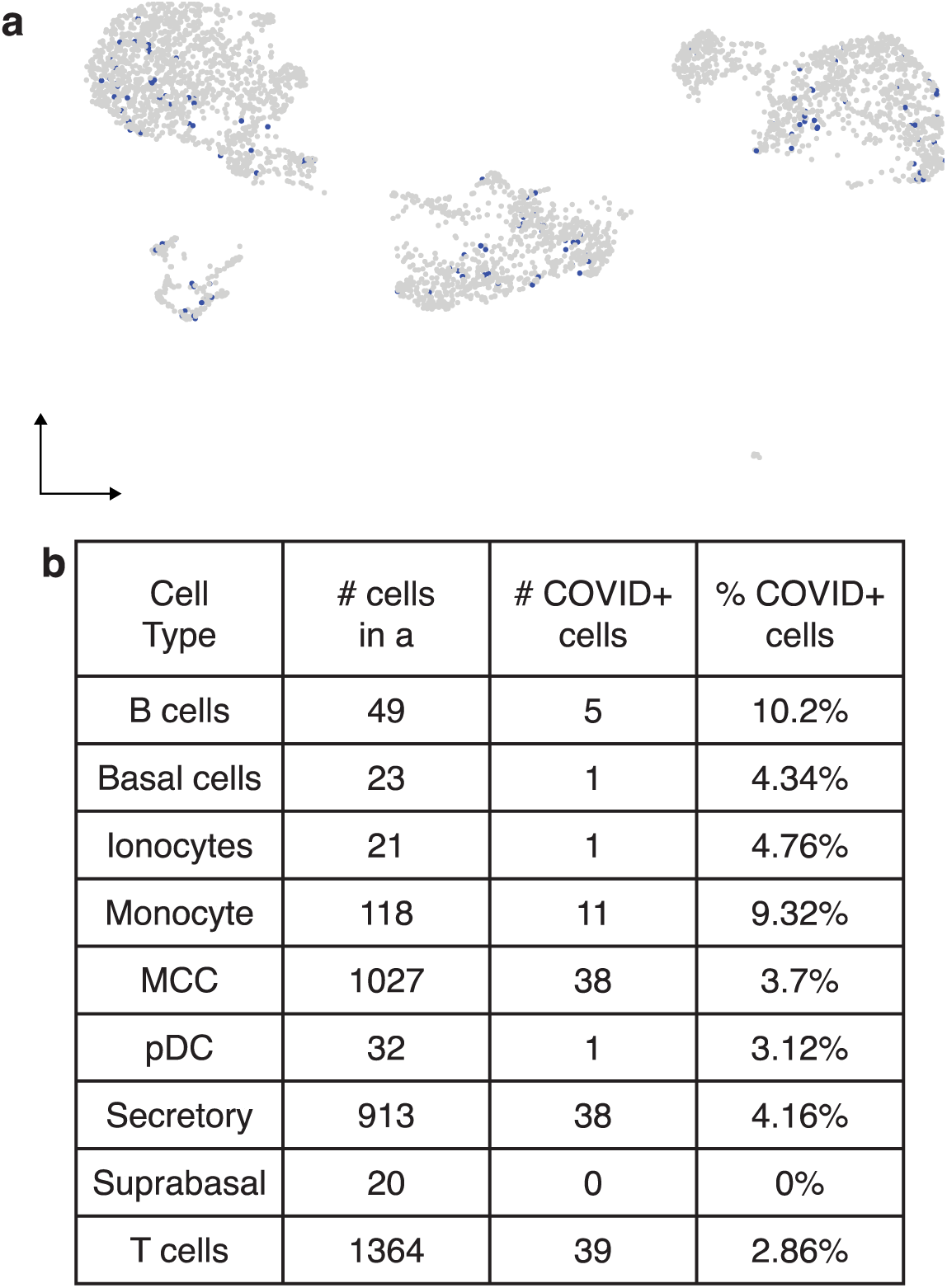
Cells in the NM of pediatric patients are susceptible to infection by SARS-CoV-2. A-B) UMAP projection of all cells from the NM samples of pediatric patients that tested positive for COVID-19 (A) or Influenza B (B). Viral transcripts were detected in cells that are colored blue, indicating active infection of those cells. C) Quantification of infected cells in A & B.

**Extended Data Figure 6:**
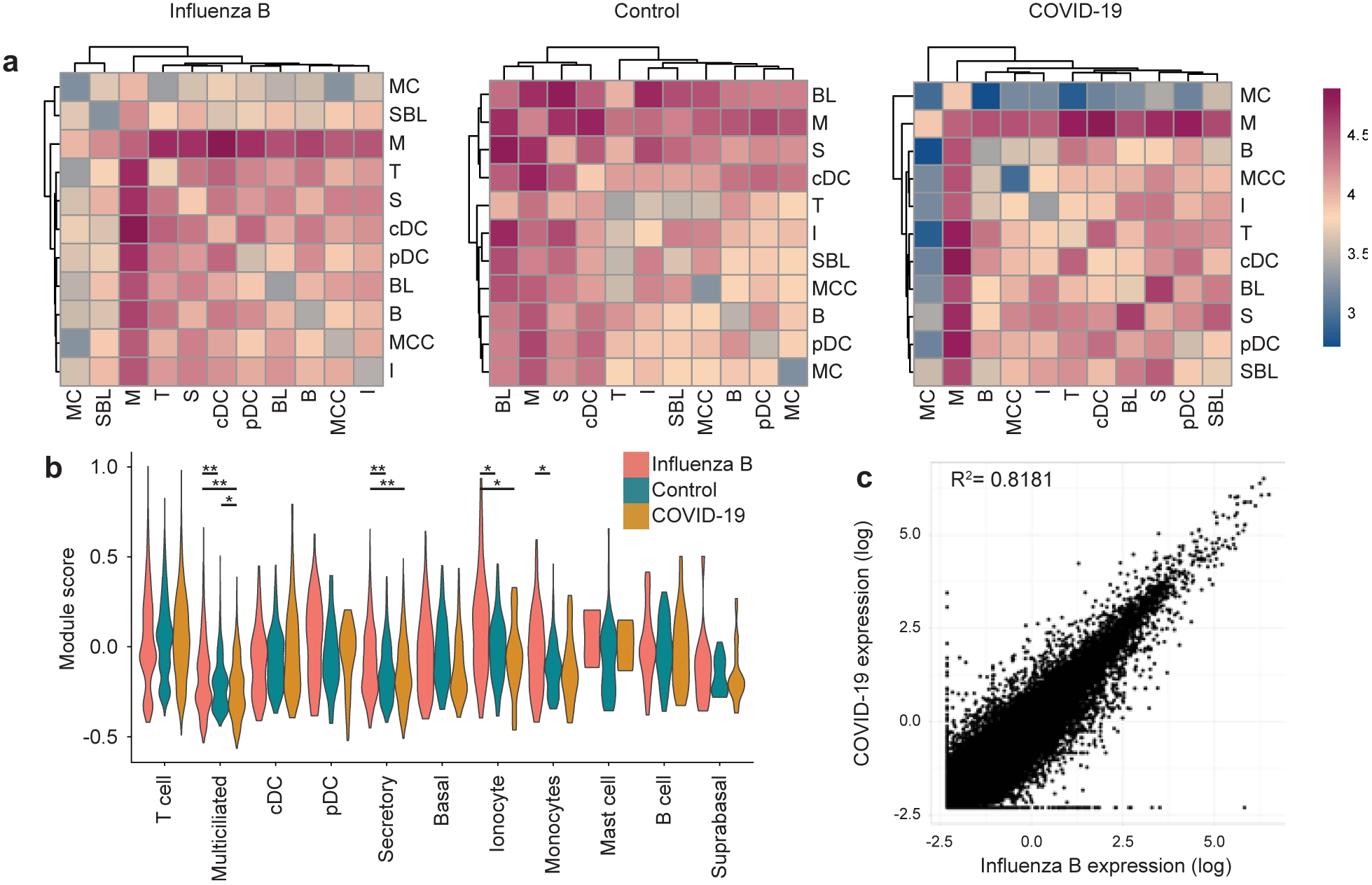
Transcriptomic response to Influenza B and COVID-19 are similar in pediatric patients. A) CellphoneDB generated heatmap of cell type interactions in Influenza B infected individuals, COVID-19 individuals, and control individuals show that cell type interactions are changed across all three viral states. Color scale is consistent across all three plots and represents the log-scaled number of ligand-receptor interaction counts between each pair of cell types. B) Violin plots of a cell death module in each cell type separated by viral state show negligible changes. p-values are for a two-sided t-test between each pairwise comparison of viral states within a cell type. All comparisons are not significant unless marked, * p-value < 0.01, ** p-value < 0.0001 C) Average scaled and normalized expression level for all genes in each cell type are similar between Influenza B (x-axis), and COVID-19 (y-axis). R^2^ shown is the Pearson’s correlation coefficient between COVID-19 expression and Influenza B expression.

**Extended Data Figure 7:**
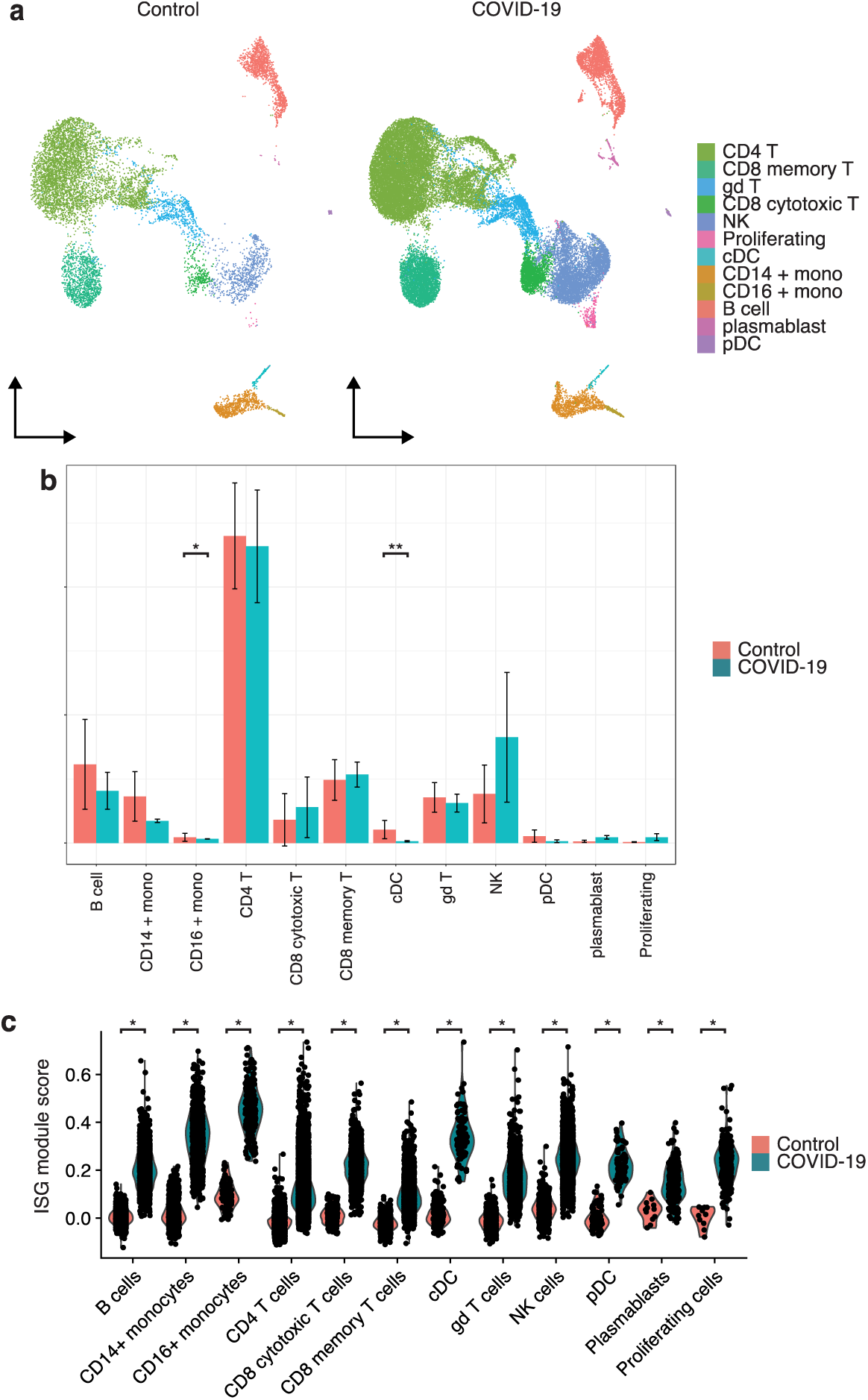
Peripheral immune response to COVID-19 in pediatric patients is similar to local response in the NM and is dominated by increased interferon signaling. A) UMAP projection of age-matched PBMCs from pediatric patients that were negative for COVID-19 (n=9) (left), or positive for COVID-19 (n=2) (right). B) Quantification of cell types present in A. Error bars show one standard deviation of the proportions present in each individual. Significance levels are for a two-sided t-test of the means of each cell type in for each individual in each viral state, all comparisons are not significant unless marked, * p < 0.05, ** p < 0.001. C) Interferon stimulated gene (ISG) module score for each cell in each cell type, separated by healthy and infected donors. Significance levels are for a two-sided t-test comparing the Control and COVID-19 donor cells within each cell type, * p-value < 0.0001.

